# Focal Adhesion Kinase Variants May Contribute to Risk of Human Myelomeningocele

**DOI:** 10.1101/2025.06.12.25329493

**Authors:** Lydia Youmans, Charani Kamath, Sara Mansoorshahi, Myra Kurjee, Parkerson Laville, Ashabari Sprenger, Jeffrey Frost, Rachel Miller, Hope Northrup, Kit Sing Au

## Abstract

Myelomeningocele (MMC) is the most severe form of an open neural tube defect (NTD) that is compatible with life. The prevalence of MMC in the United States is 1 in 2,500 live births, with the two ethnicities that have the highest occurrence of MMC being Mexican American (MA) and Caucasian American (EA). Research to date has shown that MMC results from a cumulative effect of environmental and genetic factors. Therefore, determining the underlying molecular etiology would be a step toward developing strategies for prevention and treatment.

We examined variants in 568 nervous system development genes implicated in MMC by whole exome sequencing of 254 MA and 257 EA subjects born with MMC. Mutational burden analysis was used to compare the deleterious variant load between MMC subjects and the reference population in the Genome Aggregation Exome Database (gnomADe). Higher mutational burdens were found in 18 genes, with *PTK2* being the most significant (OR=3.49, *p*=5.3e-3) among genes known to be expressed in the human neural tube at the CS12/CS13 stages. Cell migration assay was performed using seven *PTK2 (aka FAK1)* deleterious variants in transfected Fak^-/-^ mouse embryonic cells. The effect of Ptk2 knockdown on neural tube development was examined using *Xenopus* embryos.

Cell migration assay results showed the seven MMC-associated *PTK2* variants significantly affected migratory capacity compared to the wild-type *PTK2*. The Knockdown of Ptk2 significantly affected the normal neural tube closure of *Xenopus* embryos. Based on these findings, *PTK2* variants identified from MMC patients may play a role in the multifactorial causation of MMC.

## INTRODUCTION

During the 4^th^ and 5^th^ weeks of human embryogenesis, which corresponds to Carnegie stages 12 and 13, the formation and closure of the neural tube commences the development of the nervous system (https://www.ehd.org/virtual-human-embryo/about.php?stage=12). The process of neural tube formation involves an initial thickening of the dorsal ectoderm to form the neural plate. The neural plate is then induced by factors released from the notochord, a primitive mesodermal structure, to fold and fuse, forming the neural tube. The closure of the neural tube occurs in a zipper-like formation, with multiple primary sites of closure that extend cranially and caudally to assemble a long, tube-like structure. Errors in this closure process lead to the development of various forms of neural tube defects (NTDs), which are the second most common birth defects following congenital cardiovascular defects (Padmanabhan, 2006).

NTD is an overarching term that comprises a heterogeneous group of central nervous system defects. These defects can primarily affect the anterior neuropore, or closure site, causing anomalies such as anencephaly, encephalocele, or iniencephaly. They can also primarily affect the posterior neuropore, as seen in spina bifida occulta, closed spinal dysraphism, meningocele, and myelomeningocele (MMC). Both the anterior and posterior neuropores fail to close in craniorachischisis. These NTDs can also be categorized into open and closed NTDs where open NTDs (i.e., MMC) involve the protrusion of the spinal cord and neural tissue through the posterior body wall without a protective barrier, while closed NTDs (i.e., encephalocele) have skin covering over the protrusion.

MMC is the most severe form of an open NTD that is compatible with life. It accounts for more than 90% of spina bifida cases and involves the protrusion of the spinal cord and neural tissue through the posterior spine (Au, Ashley-Koch and Northrup, 2010). The prevalence of MMC in the United States is 1 out of 2,500 live births, with the two ethnicities that have the highest occurrence of MMC being Mexican Americans (MA) and Caucasian Americans (EA) in that order (Williams *et al*., 2015). In the U.S., the incidence rates of MMC are 4.17 per 10,000 in the MA population and 3.22 per 10,000 in the EA population (Boulet *et al*., 2008a), and are slowly declining, likely due to the folic acid fortification of food crops (Boulet *et al*., 2008b; Canfield *et al*., 2014). Since both these ethnicities make up a large portion of the US population, with MA being the fastest-growing ethnicity in the US, MMC poses a significant public health concern.

A more individualistic view shows that MMC also has a considerable functional toll on the patients themselves, including an inability to walk, bladder and bowel dysfunction, and associated with CNS abnormalities such as hydrocephalus and Arnold-Chiari type two malformation (Northrup and Volcik, 2000). Therefore, the management of MMC requires a multidisciplinary team that is skilled with NTDs, resulting in immense monetary burdens for medical care (Padmanabhan, 2006). The reported total healthcare cost of managing NTDs in 2013 was over $1.6 billion (Arth *et al*., 2017). Furthermore, children with MMC may have learning disabilities that require individualized educational programs to develop necessary life skills, which adds to the already hefty healthcare costs. Thus, identifying genes that correlate with susceptibility to the MMC phenotype has important implications from both public health and an economic perspective.

Numerous studies report that NTDs occur due to multifactorial causation involving environmental factors, such as hazardous chemicals, teratogens, and vitamins, as well as genetic factors (Au, Ashley-Koch and Northrup, 2010; Juriloff and Harris, 2018). So far, over a few hundred candidate genes from animal studies have been evaluated for their association with the risk of spina bifida(Hebert *et al*., 2020). These candidate genes include those involved in folate metabolism, glucose metabolism, retinoid metabolism, planar cell polarity, and apoptosis. However, of the genes studied, fewer than 20% have been shown to have even a minor effect on the risk of the spina bifida phenotype (Au, Ashley-Koch and Northrup, 2010). Landmark positive finding of folic acid supplement for NTD clinical trials (‘MRC Vitamin Study Research Group Prevention of neural tube defects: results of the Medical Research Council Vitamin Study.’, 1991) followed by finding of MTHFR genotypes associated with gene product activities (van der Put et al., 1998). These studies led to the United States Food and Drug Administration mandating the fortification of all enriched cereal grain products with folic acid in January 1998. As a result, a 22.9% decrease in the prevalence of spina bifida in the U.S. between October 1998 and December 1999, compared to the prevalence 1995-1996 (Grosse et al., 2016). The positive effect of the FA fortification experience provides promise for similar benefits from research into other genes that may correlate with the phenotype.

This study aims to examine various genes involved in nervous system development to assess association with the risk of MMC. Studies conducted with transcription factor genes involved in embryogenesis have identified some significant genes in the *PAX* family associated with NTDs (Au, Ashley-Koch and Northrup, 2010). However, further research is necessary on the genes involved in the process of neural tube formation and closure. The clinical significance of this study is utilizing the increased knowledge of associated genes to improve screening and management for MMC. This endeavor will help medical professionals set more attainable goals for MMC patient care and explore new avenues for treatment.

Genes regulating focal adhesion and cell motility play a crucial role in nervous system development (Papusheva *et al*., 2009; Navarro and Rico, 2014). We have reported many deleterious variants in cell motility (GO:0048870) genes in our MMC cohort, and these genes have been shown to contribute to NTDs in mouse models (Au *et al*., 2021). For example, Fak knockout mice showed a mesodermal deficiency, resulting in wavy neuroectoderm, involuted mesenchyme, and a deformed headfold in some embryos (Furuta *et al*., 1995; Ilić *et al*., 1995). The expression of *fak* during *X. laevis* embryo developmental stages had been established (Hens and DeSimone, 1995). Fak morphant *Xenopus* embryos express increased levels of anterior neural markers while exhibiting reciprocally reduced expression of posterior neural markers. Posterior neural plate folding and convergence-extension are also inhibited. This anteriorized phenotype resembles that of embryos knocked down zygotically for canonical Wnt signaling. FAK and Wnt3a genes are both expressed in the neural plate, and Wnt3a expression is FAK-dependent (Fonar *et al*., 2011).

In this report, we assess the deleterious variant burden among 568 protein coding genes cataloged to the biological process of nervous system development and perform experiments to reveal the functional significance of the MMC-associated variants.

## MATERIALS AND METHODS

### Study Design

Our study population is composed of subjects recruited from four different spina bifida clinics across the United States and Canada. Out of the 1,168 subjects with MMC seen in these clinics, our study population focuses on 254 MA patients with MMC and 257 EA patients with MMC since these ethnicities are most affected (https://www.cdc.gov/spina-bifida/data/index.html). Eligibility for the study was limited to patients with isolated, non-syndromic MMC at birth. The subjects that were recruited and/or their parents consented to enrollment in the study in accordance with an institutional IRB at the University of Texas Health Science Center at Houston, as described in Au et al., 2008.

The ethnically matched control group used was obtained from publicly available data on the Genome Aggregation Database (gnomAD, https://gnomad.broadinstitute.org/; Chen et al., 2024). The control population for the MA cases consists of 8,556 control Latin/Admixed American (AMR) exomes. The control population for the EA cases consists of 21,384 control Non-Finnish European (NFE) exomes.

### Exome Sequencing and Variants Calling

Whole exome sequencing (WES) from subjects and their parents was compared to WES data from an ethnically matched control population (Hillman et al., 2020).

Deleteriousness of variants was determined by predicted protein truncation (i.e., frame-shifting indels, stop-gained, and splice site motifs) and non-synonymous variants with a Combined Annotation Dependent Depletion (CADD, https://cadd.gs.washington.edu/; Rentzsch et al. 2021) score ≥20, which represents the top 1% of the most deleterious variants. Only variants with a population frequency of ≤0.001 were used in the study.

### Nervous System Development Candidate Gene List

The Gene Ontology online tool was used to acquire a list of genes involved in growth and nervous system development in *Homo sapiens* (‘The Gene Ontology Resource: 20 years and still GOing strong’, no date; Ashburner *et al*., 2000; Carbon *et al*., 2009). The keywords used for this search were “growth factor” and “nervous system development.” This search yielded a list of 573 genes that met the criteria. Of these 573 genes, we filtered the list to focus on the genes that specifically make protein products. This process resulted in a final gene list of 568 (supplementary table) genes that pertained to the gene group in question.

### Mutational Burden Test

Mutational Burden Test is performed on variants (gnomADe NFE or AMR rare allele frequency <1%) of the 568 selected genes using the method described in Hillman et al. (2020). Deleterious variants include frameshifting indels, stop-gain, stop-loss, splice motif variants, and missense variants with a CADD phred score ≥ 20 (Rentzsch *et al*., 2021). False discovery was further limited by including only variants whose positions were covered at a read depth minimum of 10 in 89.5% of both cases and reference population samples (Guo *et al*., 2018). For cases, we only counted the number of individuals carrying at least one qualifying variant within a gene. For reference population controls, the number of individuals with a qualifying variant in a gene was estimated based on the cumulative gnomAD allele frequency (AF) and gnomAD population size, assuming the Hardy-Weinberg equilibrium. We used an R package to analyze mutational burden by gene using a Fisher Exact Test, comparing the number of case individuals with and without a qualifying variant to the number of estimated reference controls individuals with and without a qualifying variant (R Core Team R, 2019). The results generated Q-Q plots for the mutational burden analysis with a lambda value greater than one were used. The genes with an odds ratio (OR) greater than one were only analyzed further. A Bonferroni correction for the number of genes tested for each network was calculated as P < 0.05/568 (8.80×10^-5^). Nominally significant genes are those with a P-value ≤ 0.05 for follow-up study.

### Cell Lines and Plasmids

*PTK2* cDNA and PTK2 variants were synthesized by subcloning downstream and in-frame of EGFP using the pcDNA3.1+N-eGFP plasmid (GenScript). Sequences of PTK2 and variants were verified by sequencing. The Fak^-/-^ mouse embryonic fibroblast cells (Ilić *et al*., 1995) are a kind gift from Dr. David D. Schlaepfer.

### Plasmid Transfection

Confluent Fak^-/-^ cells T25 flasks were trypsinized, and the detached cell concentration was measured using a hemocytometer. Approximately 250,000 cells in 1 mL of DMEM with 10% fetal bovine serum (FBS) are transferred into each well of a 24-well plate and gently rocked to distribute cells evenly. The plate was incubated overnight at 37°C in a 5% CO_2_ incubator to allow for cell attachment.

Nine of the wells were selected based on the viability and spread of the cells. The medium in the wells was discarded and replaced with 150 uL of DMEM without serum.

To prepare the transfection mixes, 0.5 ug of each plasmid was suspended in 50 uL of DMEM without serum in nine separate centrifuge tubes. In nine other tubes, 1uL of the Dreamfect (OZ Biosciences) transfection reagent was suspended in 50uL of DMEM without serum. The Dreamfect was then gently pipetted and mixed into the tubes containing the plasmids, and the combined mix was incubated at room temperature for 20 minutes.

Cells in the well plate were placed on a rotating shaker at 20 rpm and the transfection mix was added dropwise to each well. The transfected cells were incubated in a 37°C CO_2_ incubator for four hours and examined under light microscopy at hours 2 and 4 to assess cell viability. At hour 4, 250uL DMEM with serum was added to each of the nine wells, and the wells were returned to the incubator. After 30 minutes, the cells were observed under light microscopy to reaccess viability. If reduced viability, the existing medium in the wells was removed and replaced with 500 uL DMEM with serum, and the wells were incubated overnight.

The next morning, the cells were observed under a fluorescence microscope to check for eGFP expression in all nine wells. Images of the fluorescent protein in cells in the center and at the edge of the wells were obtained to account for different cell densities of each area. The same areas were observed under brightfield microscopy, and imaging was obtained to approximate the transfection efficiency.

### Cell Motility Assays

To test the effect of the deleterious variants on the functionality of PTK2 and the ability of cells to migrate, a Boyden chamber assay was conducted as previously described (Sprenger *et al*., 2023). Briefly, mouse FAK^-/-^ cells transfected with either eGFP (negative control), eGFP tagged PTK2 wild type (*wt*, positive control), or eGFP tagged PTK2 variants (S153N, N629D, P733L, I799T, R824C, E900K, and S940G) were examined for eGFP expression under fluorescence microscopy to ensure comparable transfection efficiency. Cells were trypsinized to be released from the well and were suspended in DMEM without serum before transferring into an 8 μm pore insert well (Corning Life Science) and the bottom well, containing DMEM with 10% serum. Cells were incubated in a 37°C CO_2_ incubator for 6 hours to allow the cells to pass through the pores of the insert well membrane. Cells were removed from the upper surface of the insert membrane by scrubbing with a cotton-tipped swab twice. Then, cells on the bottom membrane were fixed with 3% paraformaldehyde in PBS for 10 minutes and rinsed three times with PBS. The membrane was then excised carefully, and cells on the bottom side of the insert membrane were mounted on microscopic slides using ProLong Diamond Antifade Mountant with DAPI (ThermoFisher Scientific) for nuclear staining. Slides were observed under fluorescence microscopy at 20x magnification, and images of cells fluorescing with eGFP and DAPI were captured in 10 discrete areas. The number of fluorescent cells on the bottom surface of the insert was counted using ImageJ (https://imagej.net/ij/). Repeat experiments were performed independently by CK and MK. Two-tailed Student’s T-tests between PTK2 *wt*, variants, and plasmid control were performed.

### *Xenopus* Embryo Preparation and Morpholino-Mediated Knockdown

*X. laevis* embryos were obtained through standard beta human gonadotropic hormone-induced ovulation and *in vitro* fertilization. Embryos were cultured in 0.3 X Marc’s Modified Ringer’s (MMR) solution at room temperature and staged according to the Faber and Nieuwkoop staging system (Nieuwkoop & Faber, 1994; https://www.xenbase.org/xenbase/landmarks-table.do).

Referring to published translation-blocking morpholino oligonucleotides (MOs) targeting *X. laevis* Ptk2 (Bjerke *et al*., 2014), a Ptk2 MO5 (Xenbase.org) was selected (**Figure 1**) and synthesized (Gene Tool, LLC) for microinjection into one to two-cell stage embryos throughout the study. Control embryos were injected with a standard control MO. Each experimental group consisted of at least 30 embryos injected with 30 ng of morpholinos, and experiments were repeated three times to ensure reproducibility.

**Figure 1:**
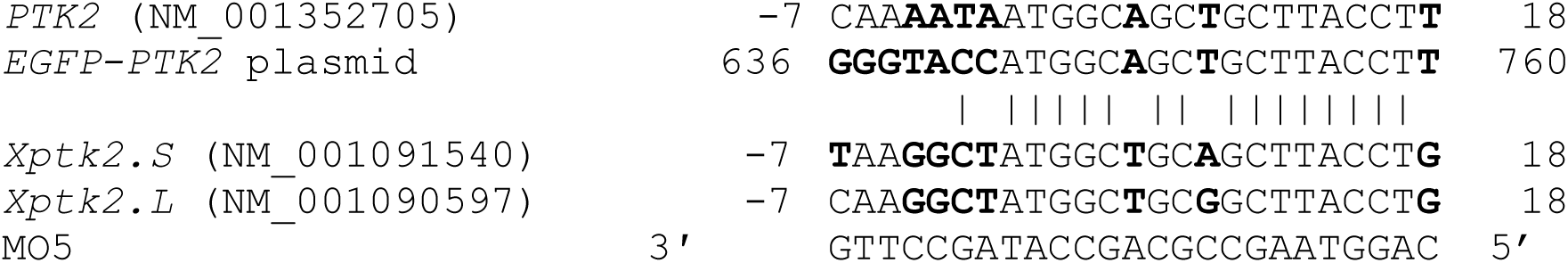
Bolded nucleotides in human *PTK2* and eGFP-*PTK2* are mismatches with *Xenopus ptk2*.

### Microinjection of Ptk2 MO and Synthesis of PTK2 mRNA

The cDNA of human *PTK2* wild-type and patient-specific variants (E900K, S940G) were cloned into expression vectors and transcribed in vitro to generate capped mRNA. A pcDNA3.1-N-EGFP plasmid is used with the eGFP ORF tagged to the N-terminus of the *wt* and variants PTK2 ORF to monitor expression and injection success. The expression of the human *PTK2* mRNA constructs using the mMESSAGE mMACHINE® T7 Transcription Kit (ThermoFisher Scientific) and translation of mRNAs in embryos was also confirmed by visualizing eGFP fluorescence under a microscope and by Western blot analysis.

### Phenotypes Analysis

Embryos were observed at two developmental stages of *Xenopus* embryos, at gastrulation and neurulation, to assess defects of gastrulation and neural tube closure. Defects were categorized as normal or abnormal, and phenotypic severity was quantified.

### Western Blot to Examine Protein Expression

Western blot analysis was conducted to validate Ptk2 knockdown using 30ng of Ptk2 MO5. The expression of human *PTK2* constructs was performed after each embryo was injected with 1-4ng of mRNA. Embryo lysates were prepared at Stage 12, and proteins were separated by SDS-PAGE, transferred to nitrocellulose membranes, and probed with antibodies against PTK2, GFP, and GAPDH (loading control).

## RESULTS

### Identify Genes with High Deleterious Variants Burden

Deleterious Variants Burden Test results among 568 genes involving nervous system development showed 18 genes with nominal significant p-value <0.05 with 11 genes known to express in CS12/CS13 neural tube (Krupp *et al*., 2012). No genes reach Bonferroni significance due to the limitation of the small sample size. In the 11 genes, *PTK2* showed the lowest nominal *p*-value of 5.33 x 10^-3^ with an OR of 3.49 (CI 1.60-7.42) (**Table 1**). Another high mutational burden gene, *CDKN1B,* with an OR=5.15 (CI 2.11-12.28, p=1.93e^-3^), was not expressed in the CS12/CS13 neural tube, prompting the follow-up assays in this study to focus on the PTK2 variants.

**Table 1:** EA-European American, MA-Mexican American, ctrls-control/reference populations in gnomAD NFE (Non-Finnish European exomes) or ARM, OR (CI) – Odd Ratio (Confident Interval). NT-neural tube expression (Krupp *et al*., 2012). with – number of individuals with deleterious variants, without-umber of individuals without deleterious variants.

| Table 1. Results of Mutational Burden Test on 563 Nervous System Development Genes |  |  |  |  |  |  |  |  |
| --- | --- | --- | --- | --- | --- | --- | --- | --- |
| Ethnicity | gene | case with | cases without | ctrls with | ctrls without | p-value | OR (CI) | NT expression |
| EA | *PTK2* | 7 | 250 | 170 | 21,214 | 5.33E-03 | 3.49 (1.60-7.42) | Yes |
| MA | *EP300* | 22 | 232 | 411 | 8,145 | 1.11E-02 | 1.88 (1.16-2.97) | Yes |
| MA | *MSX1* | 2 | 252 | 4 | 8,552 | 1.15E-02 | 16.95 (2.26-91.45) | Yes |
| MA | *EDN1* | 2 | 252 | 4 | 8,552 | 1.15E-02 | 16.95 (2.26-91.45) | Yes |
| EA | *ZFP36L1* | 3 | 254 | 39 | 21,345 | 1.35E-02 | 6.46 (1.68-19.85) | Yes |
| EA | *SIX4* | 6 | 251 | 160 | 21,224 | 1.45E-02 | 3.17 (1.35-7.28) | Yes |
| EA | *EFEMP1* | 5 | 252 | 132 | 21,252 | 2.40E-02 | 3.19 (1.23-7.81) | Yes |
| EA | *POSTN* | 19 | 238 | 963 | 20,421 | 3.39E-02 | 1.69 (1.02-2.71) | Yes |
| EA | *PLCB1* | 10 | 247 | 412 | 20,972 | 3.61E-02 | 2.06 (1.07-3.88) | Yes |
| EA | *CRKL* | 2 | 255 | 27 | 21,357 | 4.62E-02 | 6.20 (1.05-24.11) | Yes |
| EA | *EP300* | 21 | 236 | 1131 | 20,253 | 4.91E-02 | 1.59 (1.01-2.51) | yes |
| MA | *CDKN1B* | 6 | 248 | 40 | 8,516 | 1.93E-03 | 5.15 (2.11-12.28) | no |
| MA | *NCKAP1L* | 9 | 245 | 120 | 8,436 | 1.21E-02 | 2.58 (1.25-5.16) | no |
| MA | *RUNX2* | 9 | 245 | 124 | 8,432 | 1.46E-02 | 2.50 (1.21-4.98) | no |
| EA | *TAL2* | 2 | 255 | 15 | 21,369 | 1.70E-02 | 11.17 (1.82-45.70) | no |
| MA | *FUZ* | 8 | 246 | 115 | 8,441 | 2.56E-02 | 2.39 (1.06-4.86) | no |
| MA | *SOX6* | 7 | 247 | 100 | 8,456 | 3.47E-02 | 2.40 (1.09-5.24) | no |
| MA | *UBE3A* | 7 | 247 | 104 | 8,452 | 4.11E-02 | 2.30 (1.05-5.02) | no |
| MA | *MAP3K13* | 9 | 245 | 147 | 8,409 | 4.64E-02 | 2.10 (1.02-4.15) | no |

Further examination of exome data reveals a total of seven deleterious variants in nine MMC patients (7 EA and 2MA). The variants were confirmed by separate NGS experiments, which showed one of the parents carried the same variants (data not shown). The predicted functional significance is shown in **Table 2**. The locations of these variants with respect to known functional domains of PTK2 are shown in **Figure 2**.

**Figure 2:**
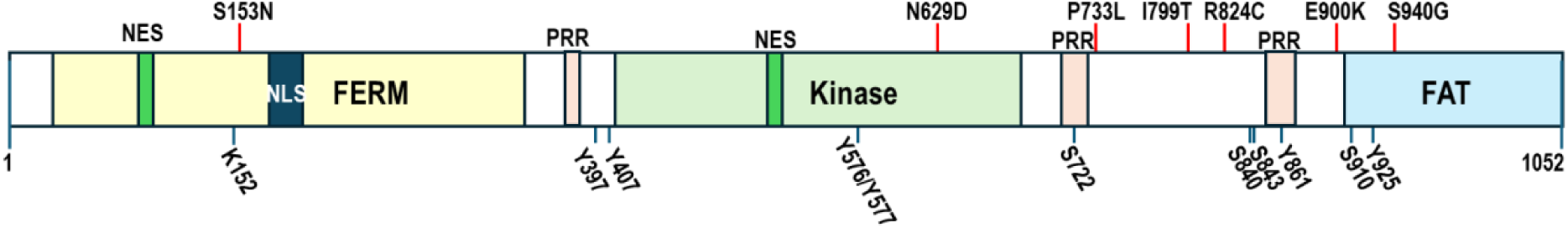
Three major domains of PTK2 are the 4.1-ezrin-radixin-moesin (FERM) domain (light yellow), kinase domain (light green), and focal adhesion targeting (FAT) domain. Other subdomains include two nuclear export signals (NES), a nuclear localization signal (NLS), and three proline-rich regions (PRR). Known phosphorylation sites (S-serine and Y-tyrosine) and a SUMOylation site K152 are shown below. Seven variants identified in MMC patients are shown in the figure above. Variant S153N is located next to the known SUMOylation site. Variant N629D is located inside the kinase domain. Variants E900K and S940G are located near or inside the FAT domain. Three other variants (P733L, I799T, and R824C are located between the second and the third PRRs.

**Table 2:** Notes: EA-European American, MA-Mexican American, NFE-Non-Finnish European exomes, AMR-Latin/Admixed American exomes. CADD-Combined Annotation Dependent Depletion, SIFT-Sorting Intolerant From Tolerant, PolyPhen-Polymorphism Phenotyping, MutPred-Mutation Prediction, PhastCons-Part of Phylogenetic Analysis with Space/Time models (Analysis of conservation by multiple sequence alignment for screening functional element).

| ethnicity | Case # | ID hg37 | NM_001352694.2 | NP_001339623.1 | gnomADe_AMR_AF | gnomADe_NFE_AF | Existing_Variation | CADD PHRED | SIFT | PolyPhen | MutPred_Top5features | MutPred_rank score | Phast Cons 100way_vertebrate | Phast Cons 100way_vertebrate_rankscore |
| --- | --- | --- | --- | --- | --- | --- | --- | --- | --- | --- | --- | --- | --- | --- |
| MA | 1 | 8:141856770:C:T | c.G458A | S153N | 0.00E+00 | 2.70E-06 | rs1429836651 | 20.4 | tolerated(1) | Benign (0.003) | Gain_of_methylation_at_K152_(P=_0.0883) | 0.48336 | 0.995 | 0.38783 |
| EA | 1 | 8:141745495:T:C | c.A1885G | N629D | 0.00E+00 | 1.80E-06 | rs879153139 | 28.9 | deleterious(0.02) | possibly_damaging (0.874) | Gain_of_ubiquitination_at_K627_(P=_0.1025) | 0.6473 | 1 | 0.71638 |
| EA | 3 | 8:141716249:G:A | c.C2198T | P733L | 1.79E-04 | 5.87E-04 | rs145314554 | 29.4 | deleterious(0) | Benign (0.175) | . | - | 1 | 0.71638 |
| EA | 1 | 8:141711093:A:G | c.T2396C | I799T | 0.00E+00 | 1.80E-06 | rs2100030036 | 18.42 | tolerated(0.58) | Benign (0.006) | Loss_of_stability_(P=_0.0081) | 0.62217 | 0.991 | 0.37257 |
| EA | 1 | 8:141711019:G:A | c.C2470T | R824C | 0.00E+00 | 6.12E-05 | rs150410292 | 34 | deleterious(0) | probably_damaging (0.996) | . | - | 1 | 0.71638 |
| MA | 1 | 8:141684408:C:T | c.G2698A | E900K | 2.32E-05 | 7.22E-06 | rs575152000 | 25.5 | tolerated(0.17) | benign(0.36) | Gain_of_ubiquitination_at_E900_(P=_4e-04) | 0.26202 | 1 | 0.71638 |
| EA | 1 | 8:141678415:T:C | c.A2818G | S940G | 9.62E-04 | 4.02E-04 | rs61733047 | 25.6 | deleterious(0.05) | possibly_damaging (0.489) | . | - | 1 | 0.71638 |

### MMC Associated PTK2 Variants Affected Cell Migration in Boyden Chamber Assays

One major function of PTK2 is modulating cell migration by facilitating focal adhesion. We set up Boyden Chamber assays to interrogate the impact of the PTK2 variants. Using FAK^-/-^ MEF cells transfected with pcDNA3.1-N-EGFP-PTK *wt*/variant, the results showed that all variants transfected FAK^-/-^ cells showed a reduction of migrating from the upper chamber to the lower chamber compared to the *wt* (**Figure 3**). The PTK2 *wt* rescued the FaK^-/-^ cells compared to the plasmid-only negative control. The effect of the N629D variant was the mildest but most significant (p=2.5e^-4^) compared to *wt*, followed by the S153N variant. All other variants (P733L, I799T, R825C, E900K, and S940G) behaved similarly to plasmid-only negative control.

**Figure 3:**
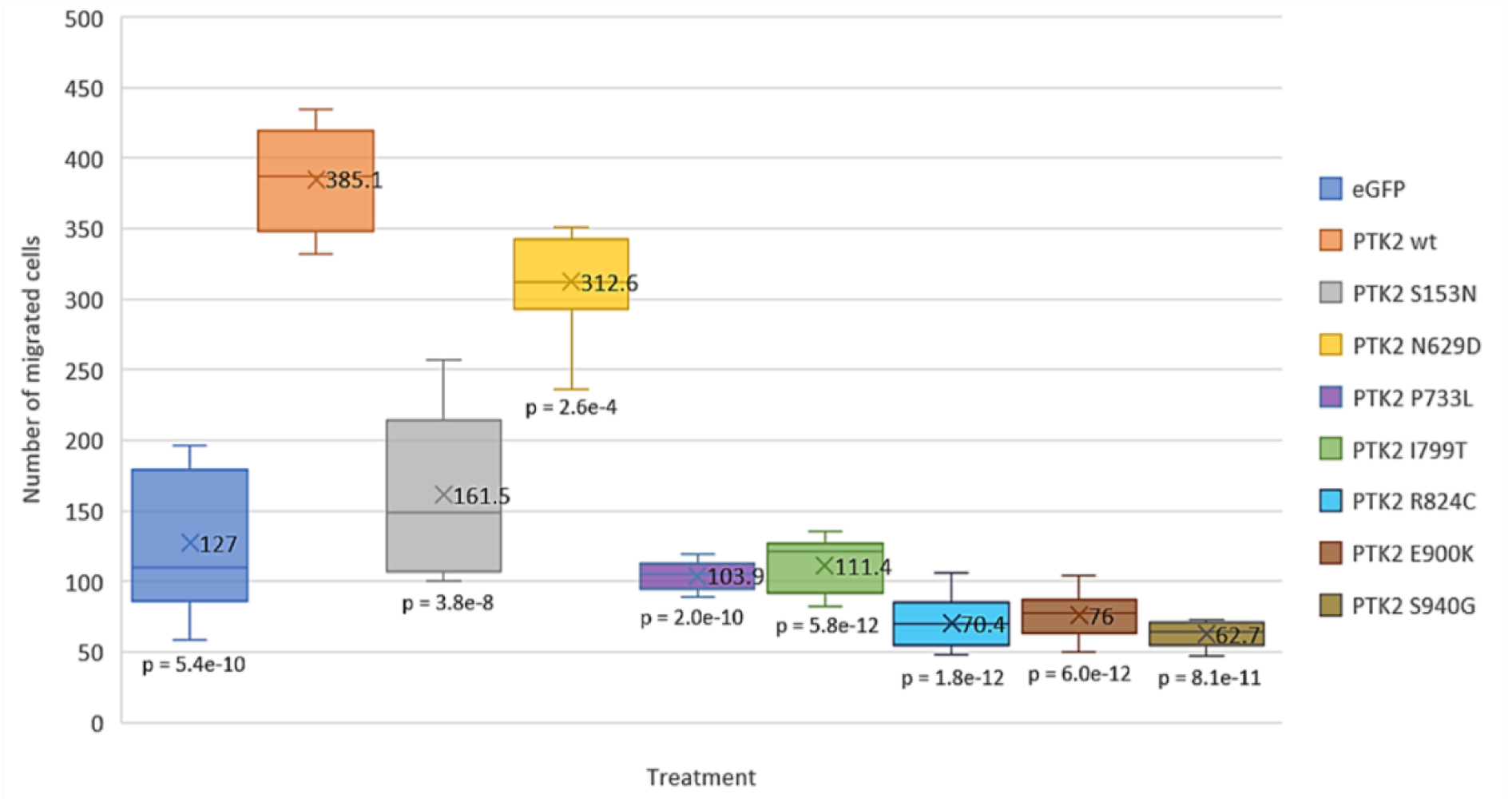
Seven MMC-associated patient variants (S153N, N629D, P733L, I799T, R824C, E900K, and S940G) of PTK2 were subcloned into a pcDNA3.1-N-eGFP-PTK2 plasmid and transfected into Fak-/-cells for Boyden chamber assays. The number of cells with eGFP signal migrated to the bottom side of the Boyden chamber membrane in 10 areas was counted, and the results are shown in boxplots. Two-tailed T-tests were performed between PTK2 *wt* and variants. The p-values were present below each box. Mean cell numbers in each of the 10 counted areas were presented and marked with X. Experiments were repeated by MK with similar observations.

### Knockdown of *X. laevis* Ptk2 Affects Gastrulation and Neural Tube Closure

To further evaluate whether PTK2 is involved in neural tube development *in vivo*, we examined the effects of Ptk2 (human PTK2 homolog) knockdown using *X. laevis* embryos. Morpholinos (**Figure 1**), targeting the *X. laevis* Ptk2 near the translation initiation codon, were designed with maximum mismatches to the *pcDNA3.1-N-eGFP-PTK2* plasmid and synthesized for the experiments to mediate knockdown of Ptk2. Results showed that the Ptk2 MO5 significantly contributed to developmental abnormalities, including both gastrulation and neural tube defects, as shown in **Figure 4a**. At the stage of the end of the gastrulation process (stage 12), Ptk2 knockdown embryos disrupted gastrulation in significant numbers (**Figure 4b**). Abnormal phenotypes included delayed closure or open blastopore lips. By the stage of the neural tube closure (stage 20), Ptk2 knockdown embryos exhibited incomplete neural tube closure (**Figure 4c**). Control embryos majorly developed normally, with normal neural tube closure phenotypes.

**Figure 4:**
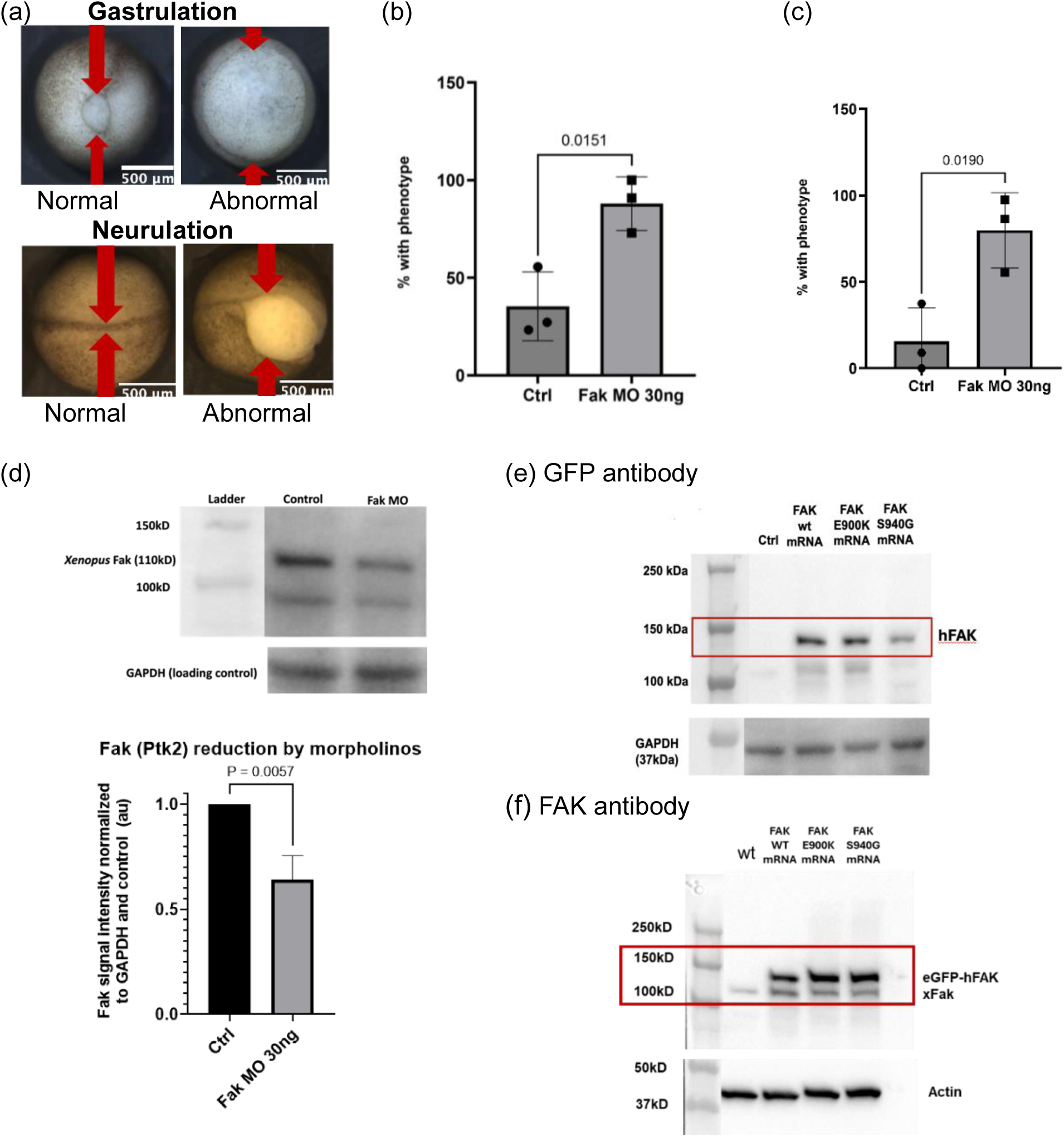
Typical gastrulation defects were observed at NF Stage 12, demonstrating delayed blastopore closure and impaired convergence-extension movements in Fak knockdown embryos compared to normal progression in control embryos. (b) Typical neurulation defects observed at NF Stage 20, characterized by failed neural tube closure and distorted dorsal morphology in Fak knockdown embryos, while control embryos display normal neural tube development. (c) Quantification of embryos exhibiting abnormal gastrulation phenotypes, comparing Fak knockdown embryos to controls injected with standard morpholino oligonucleotides (MO). (d) Western blot analysis validating the knockdown efficiency of Fak MO2 morpholinos. The blot reveals significantly reduced Fak expression in knockdown embryos compared to controls (top). Quantification of embryos with abnormal neurulation phenotypes, highlighting the increased frequency of defects in Fak knockdown embryos compared to controls (bottom). (e) Expression of human FAK (PTK2) wild-type and patient-specific variants tagged with EGFP in Xenopus embryos, confirmed using an anti-GFP antibody-bands in red box and anti-PTK2 antibody (f).

Gastrulation and neural tube defect phenotypes were common in *X. laevis* embryos after Fak knockdown. There is a significantly higher gastrulation defect (80% vs 38%, n≥30, *p=0.019) and neural tube defect (80% vs 18%, n≥30, *p=0.015) in the embryos injected with 30ng Ptk2 morpholino. The Western blot analysis confirmed efficient knockdown off Ptk2 in morpholino-injected embryos. The 110 kDa Ptk2 was significantly reduced compared to control embryos, validating the specificity and efficiency of the morpholino (**Figure 4d**).

### Full-Size EGFP-PTK2 *wt* and Variant Constructs Expression in *Xenopus* Embryos

We performed Western blots to examine the expression of human PTK2 wild-type and E900K and S940G variants in micro-injected *X. laevis* embryos. All *PTK2* constructs produced robust protein expression in *X. laevis* embryos, as evidenced by GFP and PTK2-specific antibody probing, which showed full-size140 kDa bands representing eGFP tagged human *wt* and variant constructs (**Figure 4e**).

## DISCUSSION

Through the exome sequencing of 511 MMC patients, we have identified deleterious variants in many genes and applied a deleterious variant burden test between our patients and the gnomAD reference populations to identify a short list of genes that may increase the risk to MMC (Hebert et al., 2020; Hillman et al., 2020). Among genes involved in nervous system development, we found that *PTK2* has a higher burden with seven deleterious variants in nine patients (9/511=0.0176) in the MMC cohort, suggesting that these deleterious variants may be associated with MMC risk. *PTK2* has a high intolerance to genetic variations (https://gnomad.broadinstitute.org/help/constraint), with a Loss Intolerance probability (pLI) score of 1 (o/e ratio of 0.3, CI=0.23 - 0.4). The Z score or missense changes is 3.76 with an o/e ratio of 0.69 (CI=0.65-0.74).

Among the seven deleterious variants, variant S153N is in the FERM domain (Frame *et al*., 2010). The FERM (i.e., 4.1-ezrin-radixin-moesin) domain of PTK2 regulates the binding of itself and other cellular partners to regulate the signaling of PTK2-partners and coordinates cellular responses such as cell migration, adhesion, polarization, survival, and apoptosis (Frame *et al*., 2010). Although it is located next to the K152, a known SUMOylation site, SUMO site prediction tools did not find that the variant S153N has a significant effect on K152 SUMOylation (Kadaré *et al*., 2003; Velatooru *et al*., 2021). The N629D variant located near the end of the kinase domain appears to have the least impact on cell migration. Its effect on the kinase function remains to be tested.

The remaining variants with P733L, I799T and R825C are located between the second and third PRR domains where multiple known phosphorylation targets (pS722, pS840, pS843 and pY861) located, the E900K is located slightly upstream of the pS910 site of the FAT domain, and S940G is in the FAT domain downstream of the pY925 site.

Some of these variants may affect phosphorylation of the known phosphorylation site that subsequently led to functional loss of PTK2 (aka FAK1). Phosphorylation of Y407 is required to recruit paxillin and vinculin to FAK1 and to ensure formation of focal adhesions and cell migration (Le Boeuf, Houle and Huot, 2004; Golubovskaya, 2014). VEGFR2-HSP90-RhoA-ROCK-FAK/Tyr(407) pathway in transducing the VEGF signal that leads to the assembly of focal adhesions and endothelial cell migration. Six tyrosine phosphorylation sites in focal adhesion kinase 1 (FAK1) serve to modulate FAK1 kinase activity or mediate FAK1 interaction with SH2-domain containing proteins. These are Y397, Y407, Y576, Y577, Y861 and Y925 (Mitra et al. 2005) (Mitra, Hanson and Schlaepfer, 2005). They are differentially phosphorylated by diverse agonists and implicated in transmitting different signals and effects (Ciccimaro et al. 2006, Le Boeuf et al. 2004,2006) (Le Boeuf, Houle and Huot, 2004; Ciccimaro, Hevko and Blair, 2006; Le Boeuf *et al*., 2006). Y397 is the major autophosphorylation site present upstream of the FAK kinase domain (Schaller *et al*., 1994). In response to VEGF stimulation, FAK1 is recruited and autophosphorylated at Y397. This phosphorylated tyrosine then creates a binding site for other signaling proteins that link FAK1 to downstream signaling pathways and the actin cytoskeleton (Toutant *et al*., 2002). Analysis using netphos-3.1 predicted S153, Y898, and S939 are potential targets of kinases suggesting the variants S153N, E900K and S940G may alter potential phosphorylation of these targets (**Table 3**) Blom et al., 2004, https://services.healthtech.dtu.dk ‘services’ NetPhos-3). However, experiments are needed to unravel whether these changes have significant biological consequences.

**Table 3:** Notes: Amino acid and the number in PTK2 *wt* of phosphorylation sites show the first column and bolded with the flanking sequences in the second column. Predicted score and kinase are shown in the third column and fourth column. Follow by amino acid and the number in PTK2 variants show the fifth column and bolded with the flanking sequences in the sixth column. Predicted score and kinase are shown in the last two columns. CKII-Casein Kinase II, Cam-II-Calcium/Calmodulin-Dependent Protein Kinase II, EGFR-Epidermal growth factor receptor, GSK3-Glycogen synthase kinase-3 beta, INSR-Insulin receptor, SRC-Proto-oncogene tyrosine-protein kinase Src, unsp-unspecified.

| **PTK2**<br>**wt** | **Sequence** | **Score** | **Kinase** | **PTK2**<br>**var** | **Sequence** | **Kinase** | **Score** |
| --- | --- | --- | --- | --- | --- | --- | --- |
| 153 S | QQVK**S**DYML | 0.879 | unsp | 153 N | QQVK**N**DYML | . | **0** |
| 153 S | QQVK**S**DYML | 0.531 | CKII | 153 N | QQVK**N**DYML | . | **0** |
| 153 S | QQVK**S**DYML | 0.445 | CaM-II | 153 N | QQVK**N**DYML | . | **0** |
| 153 S | QQVK**S**DYML | 0.43 | GSK3 | 153 N | QQVK**N**DYML | . | **0** |
| 397 Y | ETDDYAEII | 0.977 | unsp | 397 Y | ETDDYAEII | unsp | 0.977 |
| 397 Y | ETDDYAEII | 0.569 | INSR | 397 Y | ETDDYAEII | INSR | 0.569 |
| 397 Y | ETDDYAEII | 0.512 | EGFR | 397 Y | ETDDYAEII | EGFR | 0.512 |
| 397 Y | ETDDYAEII | 0.485 | SRC | 397 Y | ETDDYAEII | SRC | 0.485 |
| 898<br>Y/900E | PADSYNEGV | 0.533 | unsp | 898<br>Y/900K | PADSYN**K**GV | . | **0.356** |
| 898<br>Y/900E | PADSYNEGV | 0.453 | INSR | 898<br>Y/900K | PADSYN**K**GV | INSR | 0.466 |
| 925 Y | NDKVYENV**T** | 0.952 | unsp | 925 Y | NDKVYENV**T** | unsp | 0.952 |
| 925 Y | NDKVYENV**T** | 0.562 | INSR | 925 Y | NDKVYENV**T** | INSR | 0.562 |
| 925 Y | NDKVYENV**T** | 0.546 | SRC | 925 Y | NDKVYENV**T** | SRC | 0.546 |
| 925 Y | NDKVYENV**T** | 0.482 | EGFR | 925 Y | NDKVYENV**T** | EGFR | 0.482 |
| 939<br>S/940S | VIEM**S**SKI**Q** | 0.792 | unsp | 939<br>S/940G | VIEM**S**GKI**Q** | unsp | 0.861 |
| 940 S | VIEM**S**SKI**Q**P | 0.465 | CaM-II | 940 G | VIEM**S**GKI**Q**P | . | **0** |
| 940 S | VIEM**S**SKI**Q**P | 0.437 | GSK3 | 940 G | VIEM**S**GKI**Q**P | . | **0** |

The helical structures of FAT domain dimers contribute to the interaction with the LD4 domain of paxillin (https://www.rcsb.org/3d-view/1OW6/1). It has been demonstrated that the disruption of the H1/H2 hinge point amino acids around P944 opened the FAT domain, resulting in increased phosphorylation of S910 and Y925 (Kadaré *et al*., 2015). A 3D analysis showed that E900 is in a free loop preceding the first helix (H1) of the FAT domain. The S940 in H1 can play a critical role in stabilizing the alpha helix 1 (H1) of the FAT domain via bonding with I936, E937, K941, and Q943, facilitating dimerization of two PTK2 FAT domains (**Figure 5**). E900 and S940 are conserved between human PTK2 (NP_722560) and *X. laevis* Ptk2.L (NP_001084066) and Ptk2.S (NP_001085009) (Clustal Omega Multiple Sequences Alignment below), suggesting that the conserved peptide sequences potentially retain similar biological function (**Figure 6**).

**Figure 5:**
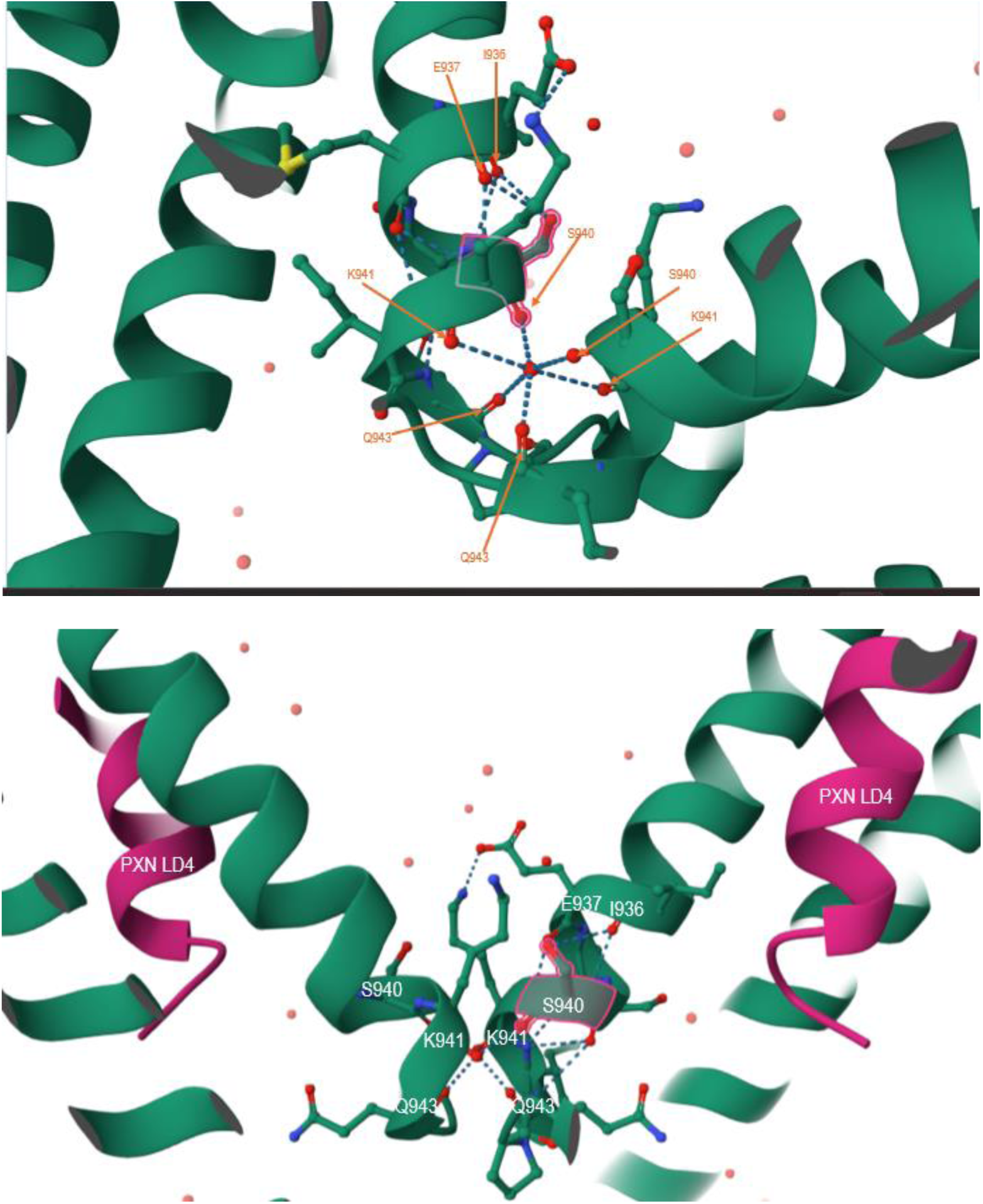
The Protein Data Base (https://www.rcsb.org/3d-view/1OW6/1) has protein 3-D data of Paxillin LD4 motif bound to the Focal Adhesion Targeting (FAT) domain of the FAK1. Zooming in to the S940 region, to review the interactions of dimers of PTK2 FAT domains and the PXN LD4 domain. S940 bonds with E936, E937 to form a stable α-helix. In addition, S940 also form H-bond with a water molecule that bond with K941 and Q943 likely stabilizing the FAT domain dimer.

**Figure 6:**
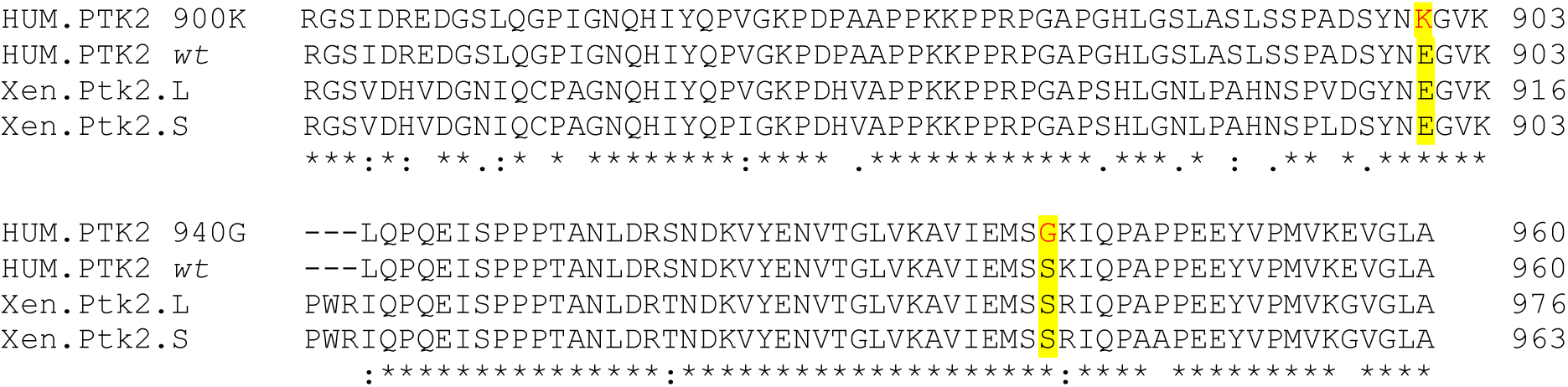
Peptide sequences of human PTK2, PTK2 variants (E900K and S949G), Ptk2.L and Ptk2.S are aligned to show the conservations between human and *X. laevis*. E900 and S940 region are well-conserved (yellow highlighted).

Through a communication with Drs. John P. Orban and Supriyo Bhattacharya, after reading their preprint on the conformational dynamics of PTK2 FAT/PXN (Bhattacharya et al., 2025; https://doi.org/10.1101/2025.01.01.630265), the S940G variant will most likely not disrupt the 4-helical bundle structure of the FAT domain without significant loss of stability. Instead, an S940G variant may increase the hydrophobicity of this part of the FAT surface. One possible effect is that the S940G variant with more intermolecular hydrophobic contacts in this region will favor PXN/FAT states, shifting the equilibrium further towards one (or more) of the four major PXN/FAT states detected in the preprint manuscript. The variant may also influence interactions with other proteins in other PTK2 signaling pathways. The higher stability of the PXN/FAT state may affect the dynamic interactions of PXN/FAT, thereby affecting the assembly and disassembly of adhesion. Stability analysis on S940G using the DUET online prediction for the effect of amino acid change on protein stability (http://biosig.unimelb.edu.au/duet/stability_prediction<u>#</u>) resulted in a decreased destabilizing effect predicted by DUET with (ΔΔG): -0.291 kcal/mol with mCSM Predicted Stability Change (ΔΔG): -0.699 kcal/mol (Destabilizing) and SDM Predicted Stability Change (ΔΔG): 0.41 kcal/mol (Stabilizing) (Pires, Ascher and Blundell, 2014). Another possibility is that S940 is a substrate of kinase(s) that has not yet been studied.

As presented in the introduction, mice with a targeted knockout of *Fak* were able to initiate gastrulation, but resulted in retarded anteroposterior axis, poor mesoderm, and involuted head mesenchyme, unable to form notochord and somite, and died between E7.5 and E10.5 (Furuta *et al*., 1995; Ilić *et al*., 1995). Heterozygotes Fak^+/-^ mice appeared normal in these studies. Other studies using conditional Fak deletion in different tissues have shown that Fak regulates the later developmental processes of vascular, neural, and cardiac tissue development (Beggs *et al*., 2003). Like the mouse models, we demonstrate that Ptk2 is critical for proper gastrulation and neural tube development in *Xenopus* embryos. Knocking down Ptk2 in *X. laevis* embryos led to increased failed gastrulation and failed neural tube closure, hinting that abnormal neurulation may be a secondary effect of abnormal gastrulation due to incomplete closure of the blastopore. Future studies will focus on evaluating the rescue effects of the wild-type and variants to elucidate further the role of PTK2 in congenital neural tube defects.

PTK2, together with Rho GTPase/GEF/ROCK, plays a key regulatory role in actin cytoskeleton organization, assembly, and disassembly upon sensing environmental signals that subsequently affect cell motility and localization of cell nuclei. Active ROCK directly phosphorylates FAK1 on S732, which triggers the phosphorylation of FAK on Y407 to modulate cell migration (Le Boeuf *et al*., 2006). Neural tube development involves the motility of neuro- and non-neuro-epithelial cells, and the formation of the median hinge point (MHP) and the dorsolateral hinge points (DLHPs) involves the movement of nuclei of some cells from a basal to apical position (McShane *et al*., 2015). Whether the functional changes of the PTK2 variants identified in our MMC patients affect the actin cytoskeleton organization, assembly, and disassembly process requires future studies to delineate.

## CONCLUSION/SUMMARY

PTK2 is a pivotal player in modulating external signals received by cells that lead to subsequent mobility reactions of cells and the localization of cell nuclei, an important process in nervous system development, especially during the early neurulation stage. A high mutation burden in *PTK2* is found in our MMC study cohort. PTK2 variants identified in MMC patients exhibited abnormal motility function, suggesting these variants may play some roles in the MMC development in these patients. Further study is warranted to delineate how these variants disrupt the neurulation process.

## Data Availability

All data produced in the present study are available upon reasonable request to the authors.

## ACKNOWLEDGEMENT

We give special thanks to Dr. David D. Schlaepfer from the Department of Reproductive Science at the University of California San Diego for gifting us the Fak^-/-^ mouse embryonic fibroblast cells.

